# Trends, Gender, and Racial Disparities in Patients with Mortality Due to Paroxysmal Tachycardia: A Nationwide Analysis from 1999-2020

**DOI:** 10.1101/2024.07.15.24310461

**Authors:** Aman Goyal, Humza Saeed, Saif Yamin, Abdullah, Wania Sultan, Muhammad Khubaib Arshad, Samia Aziz Sulaiman, Gauranga Mahalwar

**Affiliations:** Department of Internal Medicine, Seth GS Medical College and KEM Hospital, Mumbai, India; Department of Internal Medicine, Rawalpindi Medical University, Rawalpindi, Pakistan; School of Medicine, University of Jordan, Amman, Jordan; Department of Internal Medicine, Dow University of Health Sciences, Karachi, Pakistan; Cleveland Clinic Foundation, Cleveland, Ohio, USA

**Author notes:** Address for correspondence, Gauranga Mahalwar, MD, Department of Internal Medicine, Cleveland Clinic Foundation, Cleveland, Ohio, USA. **Funding:** The authors did not receive any funding to conduct the study. **Ethical considerations:** No ethical approval was required for this study design, as all data were obtained from publicly available sources. **Data Availability:** The data supporting the findings of this study are openly available in CDC Wonder at [https://wonder.cdc.gov/].

**Keywords:** Mortality, Paroxysmal Tachycardia, Epidemiology, Trends, Racial disparity, Gender disparity, Cardiology

## Abstract

**Background:** Paroxysmal tachycardia encompasses various heart rhythm disorders that cause rapid heart rates. Its episodic occurrence makes it difficult to identify and measure its prevalence and trends in the population. Additionally, there is limited data on disparities and trends in mortality due to paroxysmal tachycardia, which is essential for assessing current medical approaches and identifying at-risk populations.

**Methods:** Our study examined death certificates from 1999 to 2020 using the CDC WONDER Database to identify deaths caused by paroxysmal tachycardia in individuals aged 25 and older, using the ICD-10 code I47. Age-adjusted mortality rates (AAMRs) and annual percent changes (APC) were calculated by year, gender, age group, race/ethnicity, geographic location, and urbanization status.

**Results:** Between 1999 and 2020, 155,320 deaths were reported in patients with paroxysmal tachycardia. Overall, AAMR decreased from 4.8 to 3.7 per 100,000 population between 1999 and 2020, despite showing a significant increase from 2014 to 2020 (APC: 4.33; 95% CI: 3.53 to 5.56). Men had consistently higher AAMRs than women (4.7 vs. 2.2). Furthermore, we found that AAMRs were highest among Non-Hispanic (NH) Black or African Americans and lowest in NH Asian or Pacific Islanders (4 vs. 1.9). Nonmetropolitan areas had higher AAMRs than metropolitan areas (3.6 vs. 3.2).

**Conclusions:** Our analysis showed a significant decrease in mortality from paroxysmal tachycardia since 1999, although there has been a slight increase in recent years. However, disparities remain, with higher AAMRs among men, NH Black or African Americans, and residents of non-metropolitan areas. These findings call for immediate public health actions to curb the rising trends and reduce potential disparities.

**Clinical Perspectives What is New?:**

- In this analysis of population-level US mortality data from 1999 to 2020, we observed an overall decrease in mortality due to Paroxysmal Tachycardia, despite a significant upward trend from 2014 to 2020.
- Older adults had higher age-adjusted mortality rates than young and middle-aged adults, and men had higher mortality rates than women.
- Non-Hispanic Black or African Americans had the highest mortality rates among racial groups, and those in non-metropolitan areas had higher rates than those in metropolitan areas.

**What Are the Clinical Implications?:**

- By addressing the effects of the pandemic and racial disparities, healthcare providers and policymakers can develop effective strategies to address this concerning trend.
- Education should not only identify risk factors but also work to change the socio-cultural conditions that cause these risks.

## Introduction

Paroxysmal tachycardia, also known as episodic tachycardia, encompasses a subset of abnormal heart rhythms, including re-entry ventricular arrhythmia, supraventricular tachycardia (SVT), and ventricular tachycardia (VT). Mismanagement of any of these conditions can have lethal consequences. Among them, re-entrant arrhythmia has been associated with relatively low mortality rates. However, it still has been found to pose long-term health risks, including stroke and thromboembolism in specific subtypes (1–3).

SVT has been linked to a higher incidence in structural heart disease (SHD), occurring in up to 32% of the SHD population (3). Therefore, due to its association with SHD, it is considered the most common arrhythmia in children, affecting around 1 in 250 otherwise healthy children (2, 3). Moreover, a large multicenter study revealed that 82% of deaths of SHD-patients occurred due to SVTs (4). Simultaneously, VT posed a substantial risk of mortality and ranked as the fifth most frequent cause of emergency department visits among patients between 65 to 84 years of age (5). Moreover, it was also found that 30% to 75% of out-of-hospital cardiac arrests were attributed to VTs, adding to its dangerous nature (6).

Despite the relatively favorable patient outcomes associated with paroxysmal tachycardia, tachyarrhythmias significantly affect healthcare systems and survival rates, and have been closely tied to an increased risk of mortality, comorbidities, and complex cardiac defects (7). As far as medical management is concerned, although implantable cardioverter defibrillators have demonstrated efficacy in reducing VT-related deaths, it is revealed that they unfortunately do not mitigate the risk of VT recurrences (8).

Hence, it is imperative to explore the demographic and regional mortality patterns among US adults with paroxysmal tachycardia. By exploring the trends, gender, and racial disparities in mortality due to paroxysmal tachycardia, our study aims to identify potentially vulnerable populations to guide future research and clinical practice to improve prevention strategies and develop smarter frameworks to reduce complications associated with this condition. Additionally, our research will contribute to better healthcare outcomes and improved quality of life for affected individuals.

## Methods

### Study Setting and Population

We obtained mortality data from the Centers for Disease Control and Prevention’s WONDER database, which provides comprehensive epidemiological information. Our investigation focused on mortality rates among individuals affected by Paroxysmal Tachycardia between 1999 and 2020. Using the International Statistical Classification of Diseases and Related Health Problems, 10th Revision, we assigned code I47 to Paroxysmal Tachycardia, which inckuded 147.0, 147.1, 147.2, and 147.9 for Re-entry ventricular arrhythmia, Supraventricular tachycardia, Ventricular tachycardia, and unspecified Paroxysmal tachycardia, respectively. Our analysis centered on death certificates within the Multiple Cause of Death Public Use dataset, allowing us to explore mortality associated with these conditions in patients aged 25 years or older. Institutional review board approval was unnecessary, as we utilized a de-identified government-provided public-use dataset in accordance with Strengthening the Reporting of Observational Studies in Epidemiology (STROBE) guidelines (9).

### Data Abstraction

Our investigation analyzed demographic variables, including population size, age distribution, gender composition, racial and ethnic background, geographic location, urbanization level, and place of death. The locations of death spanned diverse settings, including inpatient facilities, outpatient clinics, emergency rooms, sudden death cases, residences, hospice/nursing homes, long-term care facilities, and instances where the location remained unspecified. We meticulously defined racial and ethnic categories, encompassing Hispanic (Latino), Non-Hispanic (NH) White, NH Black/African American, NH American Indian/Alaskan Native, and NH Asian.

In accordance with the age-wise analysis, we categorized patients into ten-year intervals, distinguishing young adults (25-44 years), middle-aged adults (45-64 years), and older individuals (65-85+ years) (10). We utilized the Urban-Rural Classification Scheme from the National Center for Health Statistics to classify our study population geographically. Urban areas housed populations of 50,000 or more, while Rural areas included locales with fewer than 50,000 residents. Furthermore, we divided the United States into four regions based on the US Census Bureau’s classification: Northeast, Midwest, South, and West (11).

### Statistical Analysis

We systematically investigated gender, race, age, urbanization, and census-related patterns by computing both crude and age-adjusted mortality rates (AAMR) per 100,000 individuals. The 2000 US population served as the reference for standardizing AAMR. To assess temporal changes in mortality rates, we utilized the Joinpoint Regression Program (Version 5.0.2, National Cancer Institute) (12). This analytical approach involved fitting log-linear regression models to the raw data trends, allowing us to calculate the annual percent change (APC) in AAMR along with its 95% confidence interval (CI). We analyzed whether APCs indicated increasing or decreasing trends by assessing their statistical deviation from the null hypothesis of zero change. Statistical significance was determined using a two-tailed t-test with a threshold of P < 0.05.

## Results

Between 1999 and 2020, there were 155,320 deaths where Paroxysmal Tachycardia was either the underlying or contributing cause (Supplemental Table 1). Place of death was recorded for 152,390 cases: 75.7% in medical facilities, 13.3% at decedents’ homes, 7.7% in nursing homes/long term care, and 1.4% in hospices. (Supplemental Table 2).

### Demographic trends in mortality

The AAMR was 4.8 (95% CI: 4.7 to 4.9) in 1999 and decreased to 3.7 (95% CI: 3.2 to 3.3) in 2020. The overall AAMR significantly decreased from 1999 to 2007 (APC: -6.24; 95% CI: -6.82 to - 5.78), followed by a slight decrease from 2007 to 2014 (APC: -0.22; 95% CI: -1.34 to 0.85) and a significant increase from 2014 to 2020 (APC: 4.33; 95% CI: 3.53 to 5.56). (Figure 1, Supplemental Table 3 and 4)

**Figure 1.**
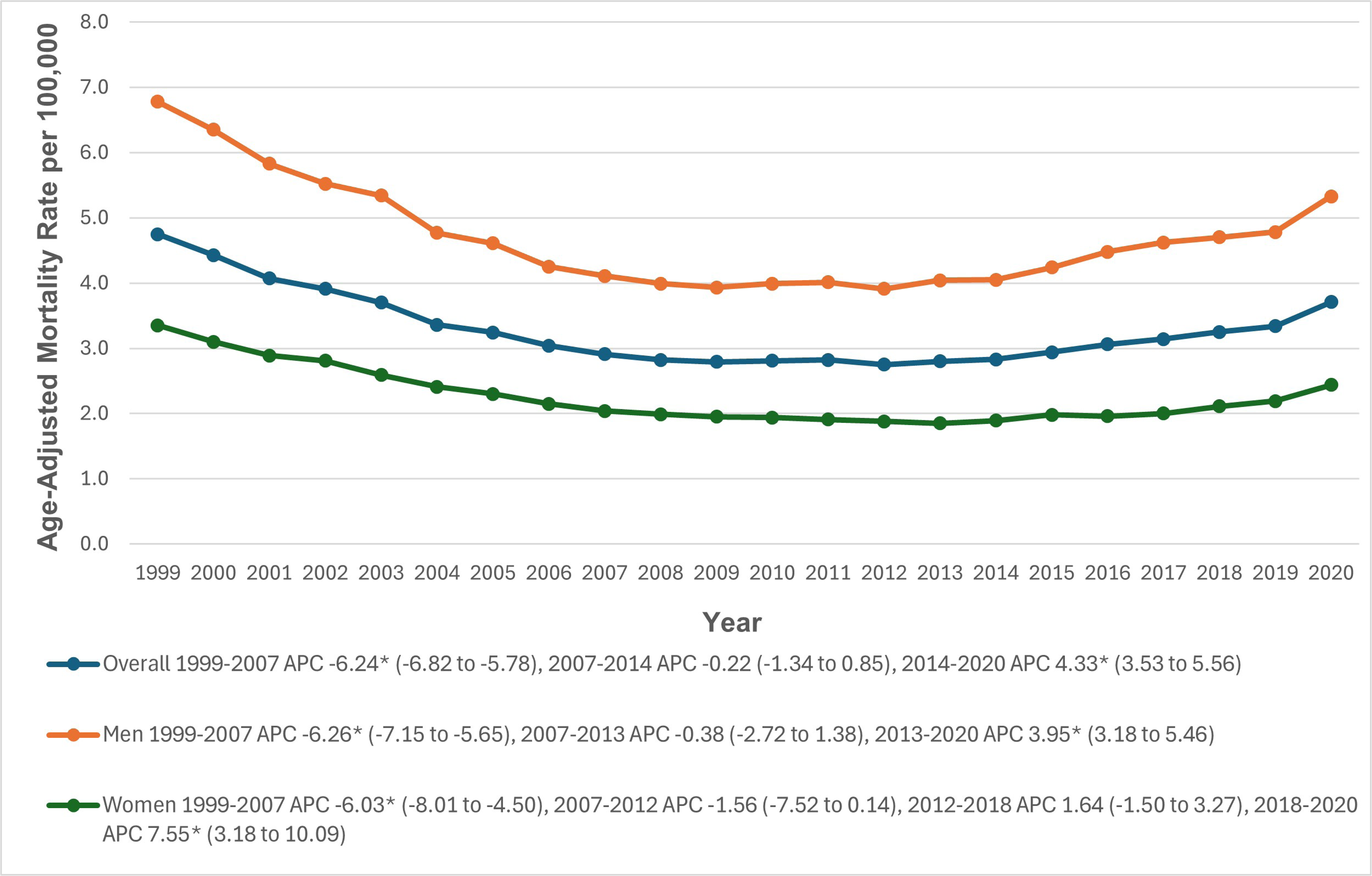
Overall and Sex-Stratified Paroxysmal Tachycardia-related Age-Adjusted Mortality Rates per 100,000 in Adults in the United States, 1999 to 2020. * Indicates that the annual percentage change (APC) is significantly different from zero at α = 0.05. AAMR = age-adjusted mortality rate.

### Gender stratification

During the study period, men’s AAMRs were higher than women’s (Men: 4.7; 95% CI: 4.6 to 4.7; Women: 2.2; 95% CI: 2.2 to 2.2). In 1999, the average AAMR for men was 6.8 (95% CI: 6.6 to 7.0), which decreased significantly to 4.1 (95% CI: 4.0 to 4.2) in 2007 (APC: -6.26; 95% CI: -7.15 to -5.65). It remained somewhat stable till 2013 (APC: -0.38; 95% CI: -2.72 to 1.38), followed by a significant increase to 5.3 in 2020 (APC: 3.95; 95% CI: 3.18 to 5.46). For women, the AAMR was 3.4 (95% CI: 3.2 to 3.5) in 1999, significantly dropping to 2.0 (95% CI: 2.0 to 2.1) in 2007 (APC: -6.03; 95% CI: -8.01 to -4.50) followed by a slight decrease to 1.9 in 2012 (APC: -1.56; 95% CI: -7.52 to 0.14). It then increased slightly to 2.1 in 2018 (APC: 1.64; 95% CI: -1.50 to 3.27), followed by a significant increase to 2.4 in 2020 (APC: 7.55; 95% CI: 3.18 to 10.09) (Figure 1, Supplemental Table 3 and 4)

### Stratification By Age groups

3When stratified by age groups, older adults had the highest AAMRs (13.3; 95% CI: 13.2 to 13.3), followed by middle-aged adults (1.7; 95% CI: 1.6 to 1.7) and young adults (0.2; 95% CI: 0.2 to 0.2). Among young adults, AAMRs significantly decreased from 1999 to 2001 (APC: -17.28; 95% CI: -24.49 to -4.77). Middle-aged and older adults observed a similar trend from 1999 to 2005 (APC: -7.27; 95% CI: -11.06 to -5.66) and from 1999 to 2007 (APC: -6.09; 95% CI: -6.78 to -5.61) respectively. From 2001 to 2013, young adults had a stable AAMR (APC: 0.22; 95% CI: -1.81 to 2.44), followed by a significant increase till 2020 (APC: 6.60; 95% CI: 3.53 to 16.37). The AAMR decreased slightly from 2005 to 2012 in middle aged (APC: -1.60; 95% CI: -3.51 to 1.86) and from 2007 to 2014 in the older adults (APC: -0.80; 95% CI: -2.43 to 0.50). This was followed by a significant increase till 2020 among both middle aged (APC: 6.40; 95% CI: 5.35 to 7.98) and older adults (APC: 3.70; 95% CI: 2.66 to 5.64) (Figure 2, Supplemental Table 3 and 5)

**Figure 2.**
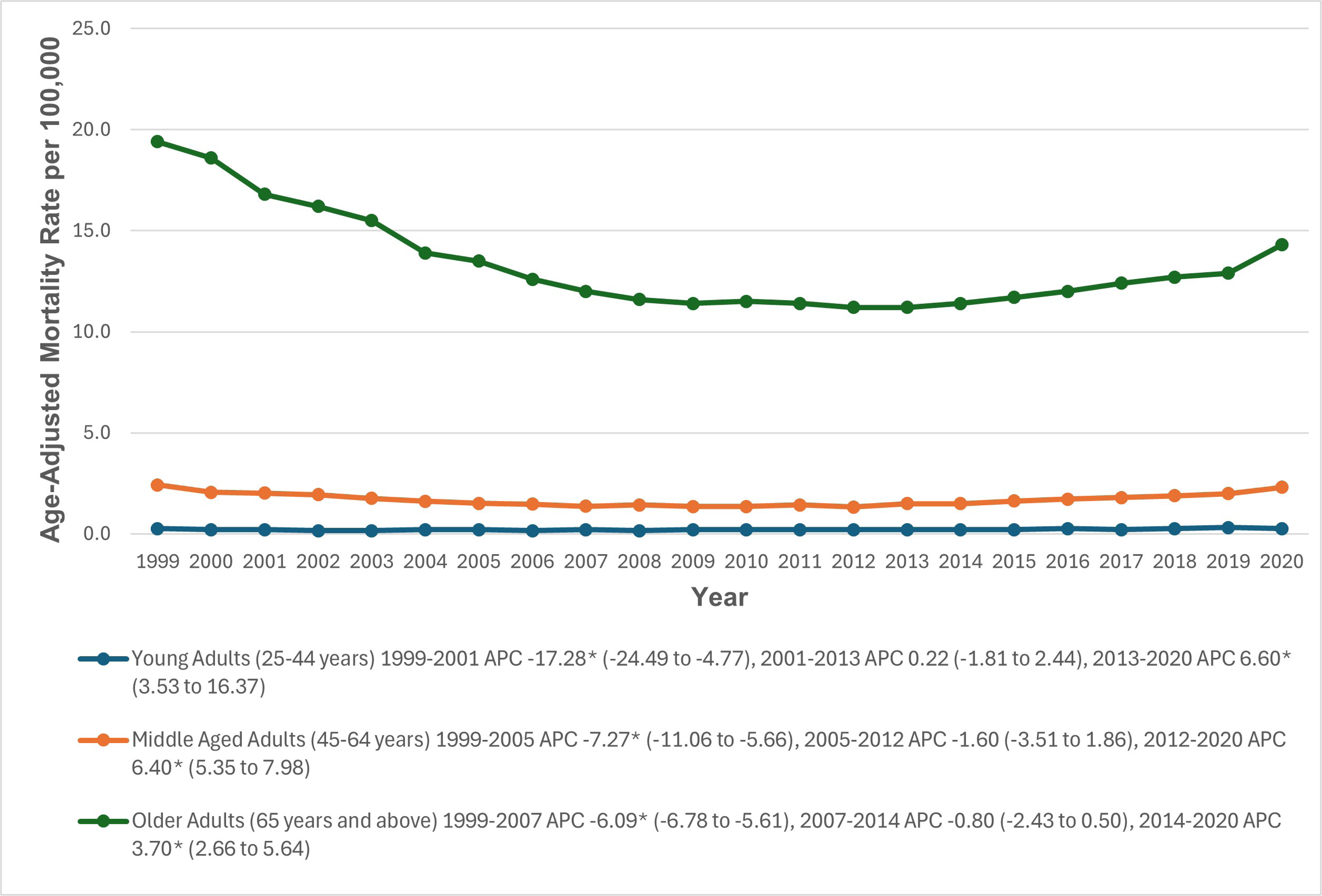
Paroxysmal Tachycardia-related Age-Adjusted Mortality Rates per 100,000, Stratified by age groups in Adults in the United States, 1999 to 2020 * Indicates that the annual percentage change (APC) is significantly different from zero at α = 0.05. AAMR = age-adjusted mortality rate.

### Racial stratification

When stratified by race or ethnicity, AAMRs were highest among NH Black or African American patients (4.0; 95% CI: 3.9 to 4.1), followed by NH American Indian or Alaska Native (3.3; 95% CI: 3.0 to 3.5), NH White (3.3; 95% CI: 3.3 to 3.4), Hispanic (2.1; 95% CI: 2.1 to 2.2), and NH Asian or Pacific Islander populations (1.9; 95% CI: 1.8 to 1.9). From 1999 to 2014, AAMRs for NH American Indians or Alaska Natives slightly decreased (APC; -2.24; 95% CI: -17.81 to 1.37), followed by a significant increase till 2020 (APC: 8.40; 95% CI: 0.53 to 33.72). Similarly, Hispanics had a significant decrease in AAMRs from 1999 to 2010 (APC: -7.01; 95% CI: -9.12 to - 5.04), followed by a significant increase till 2020 (APC: 5.21; 95% CI: 3.45 to 8.09). Among the NH Black orAfrican Americans, the AAMRs significantly decreased from 1999 to 2008 (APC: - 5.55; 95% CI: -8.68 to -4.15), followed by a slight increase from 2008 to 2016 (APC: 1.00; 95% CI: -4.60 to 2.87), and a significant increase till 2020 (APC: 6.52; 95% CI: 3.11 to 12.90). Similarly, AAMRs of NH Asians decreased from 1999-2002 (APC: -3.31; 95% CI: -9.11 to 5.52) and from 2002 to 2006 (APC: -13.31; 95% CI: -19.27 to 6.23). This was followed by an upward trend from 2006-2009 (APC: 10.11; 95% CI: -4.42 to 15.55), a decrease from 2009-2014 (APC: -4.36; 95% CI: -9.89 to 2.62) and a significant increase till 2020 (APC: 6.45; 95% CI: 4.48 to 10.44). Among the NH Whites, there was a significant decrease in AAMRs from 1999-2006 (APC: -6.05; 95% CI: - 7.57 to -5.39) and a slight decrease from 2006-2010 (APC: -2.76; 95% CI: -5.07 to 0.71). This was followed by a significant increase from 2010-2018 (APC: 2.05; 95% CI: 0.87 to 2.89) and from 2018-2020 (APC: 7.69; 95% CI: 4.42 to 9.67) (Figure 3, Supplemental Table 3 and 6).

**Figure 3.**
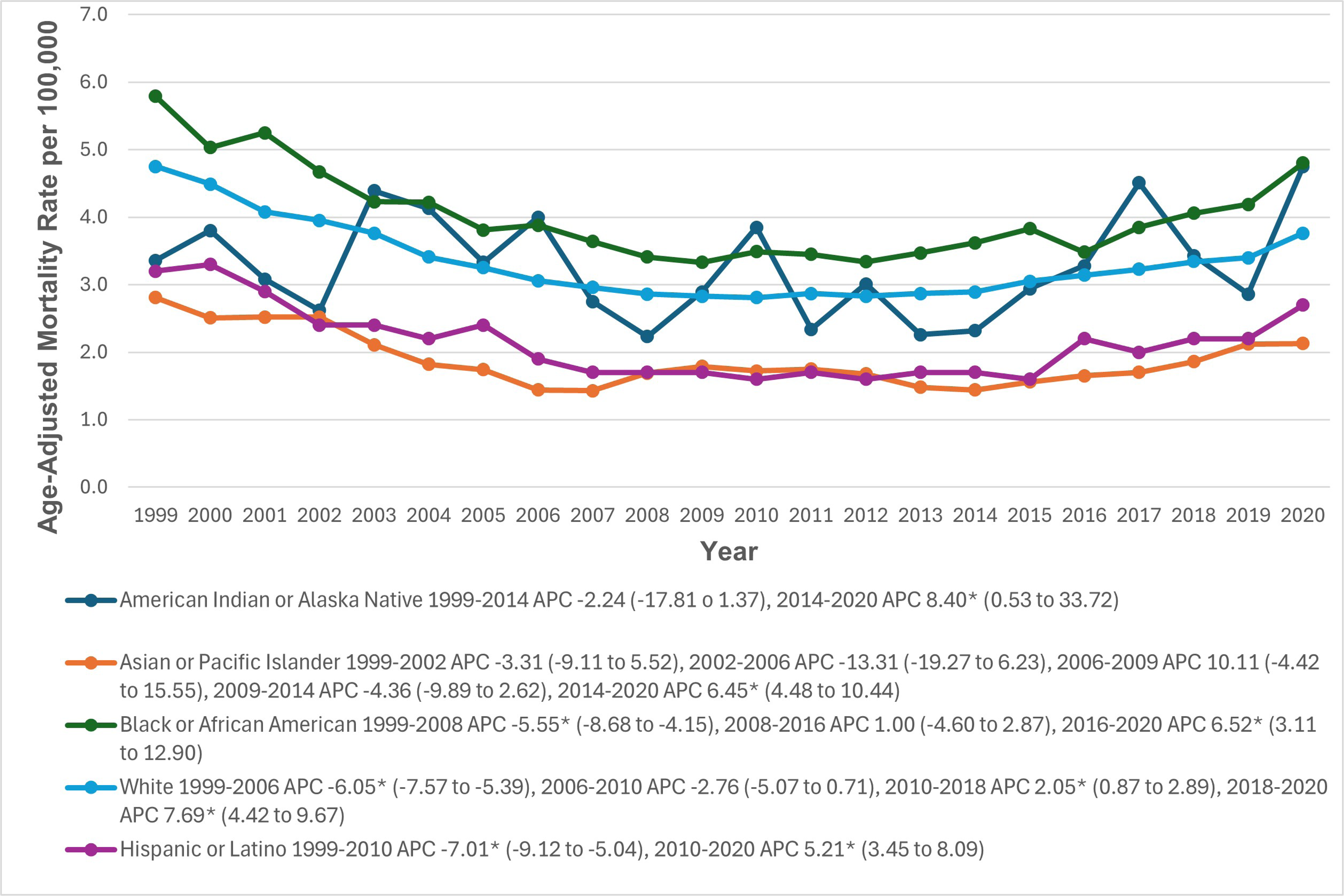
Paroxysmal Tachycardia-related Age-Adjusted Mortality Rates per 100,000, Stratified by race in Adults in the United States, 1999 to 2020 * Indicates that the annual percentage change (APC) is significantly different from zero at α = 0.05. AAMR = age-adjusted mortality rate.

### State-wise distribution

AAMR values varied significantly by state, ranging from 2.2 (95% CI: 2.1 to 2.2) in New York to 4.7 (95% CI: 4.5 to 4.9) in West Virginia. States in the top 90th percentile (Pennsylvania, Ohio, Indiana, Tennessee, South Carolina, West Virginia) had AAMRs about twice as high as those in the bottom 10th percentile (New York, Louisiana, New Mexico, Florida, Arizona, Massachusetts, Minnesota) (Figure 4, Supplemental Table 7)

**Figure 4.**
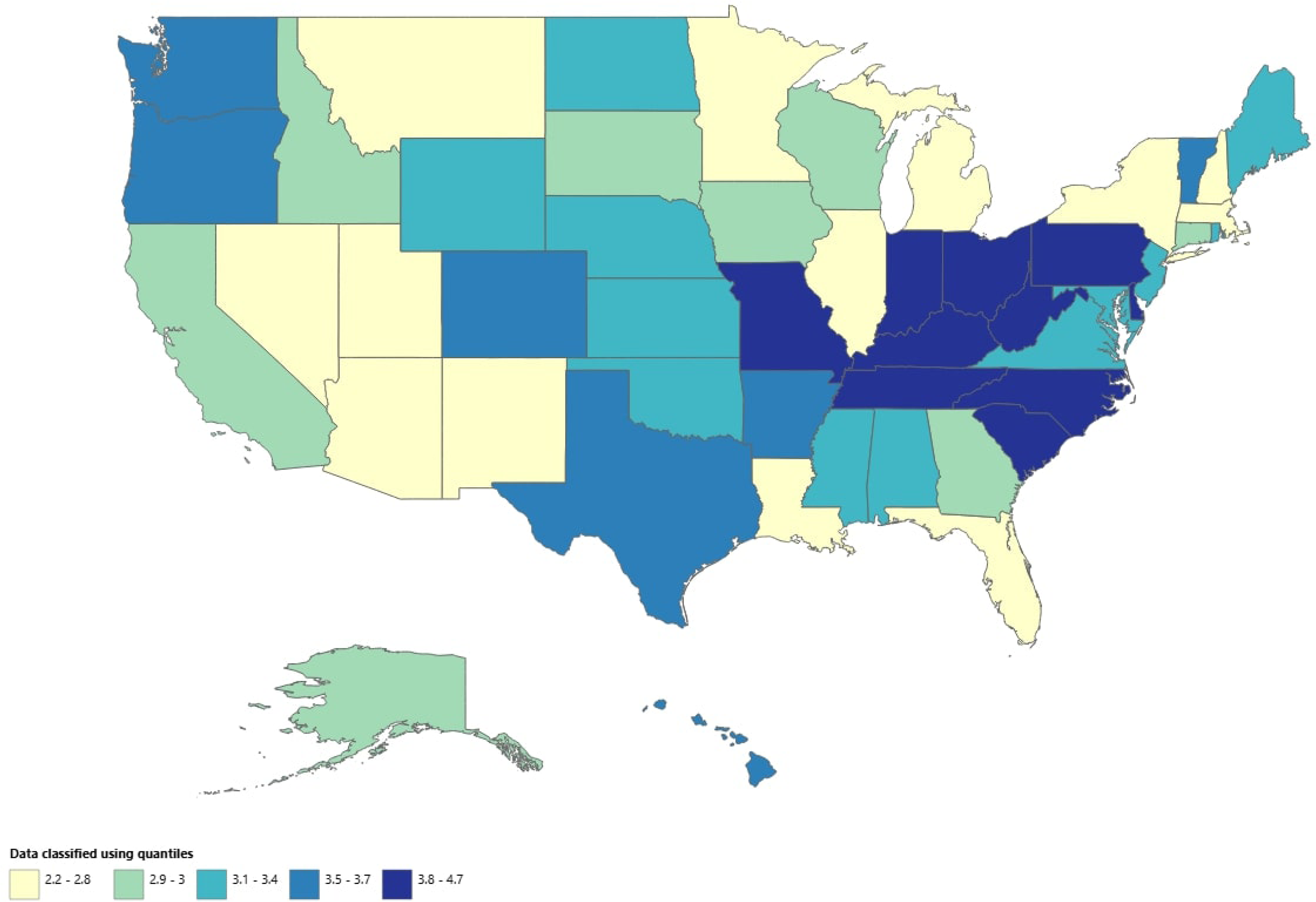
Paroxysmal Tachycardia-related Age-Adjusted Mortality Rates per 100,000, Stratified by State in Adults in the United States, 1999 to 2020.

### Census Region

During the study period, the AAMRs differed slightly across the census regions, with the highest in midwestern and southern regions (3.4; 95% CI: 3.4 to 3.4), followed by Western (3.1; 95% CI: 3.0 to 3.1), and Northeastern (3.0; 95% CI: 3.0 to 3.1) regions. From 1999 to 2009, AAMRs decreased significantly in the midwestern region (APC: -5.85; 95% CI: -6.68 to -5.18), followed by a significant increase till 2020 (APC: 2.91; 95% CI: 2.18 to 3.83). In the southern region, AAMRs decreased significantly from 1999 to 2005 (APC: -6.73; 95% CI: -8.19 to -5.93), followed by another significant decrease from 2005 to 2012 (APC: -2.55; 95% CI: -3.89 to -0.79) and a significant increase till 2020 (APC: 3.45; 95% CI: 2.73 to 4.47). The western region had similar trends with an initial significant decrease from 1999 to 2006 (APC: -5.21; 95% CI: -9.47 to -3.65), followed by a slight decrease from 2006 to 2013 (APC: -1.41; 95% CI: -3.90 to 3.97) and a significant increase till 2020 (APC: 5.20; 95% CI: 3.29 to 10.88). The northeastern region initially observed a significant decrease from 1999 to 2007 (APC: -7.48; 95% CI: -8.42 to -6.89), followed by a significant increase from 2007 to 2018 (APC: 1.01; 95% CI: 0.10 to 1.75) and from 2018 to 2020 (APC: 11.50; 95% CI: 5.78 to 14.42) (Figure 5, Supplemental Table 3 and 8)

**Figure 5.**
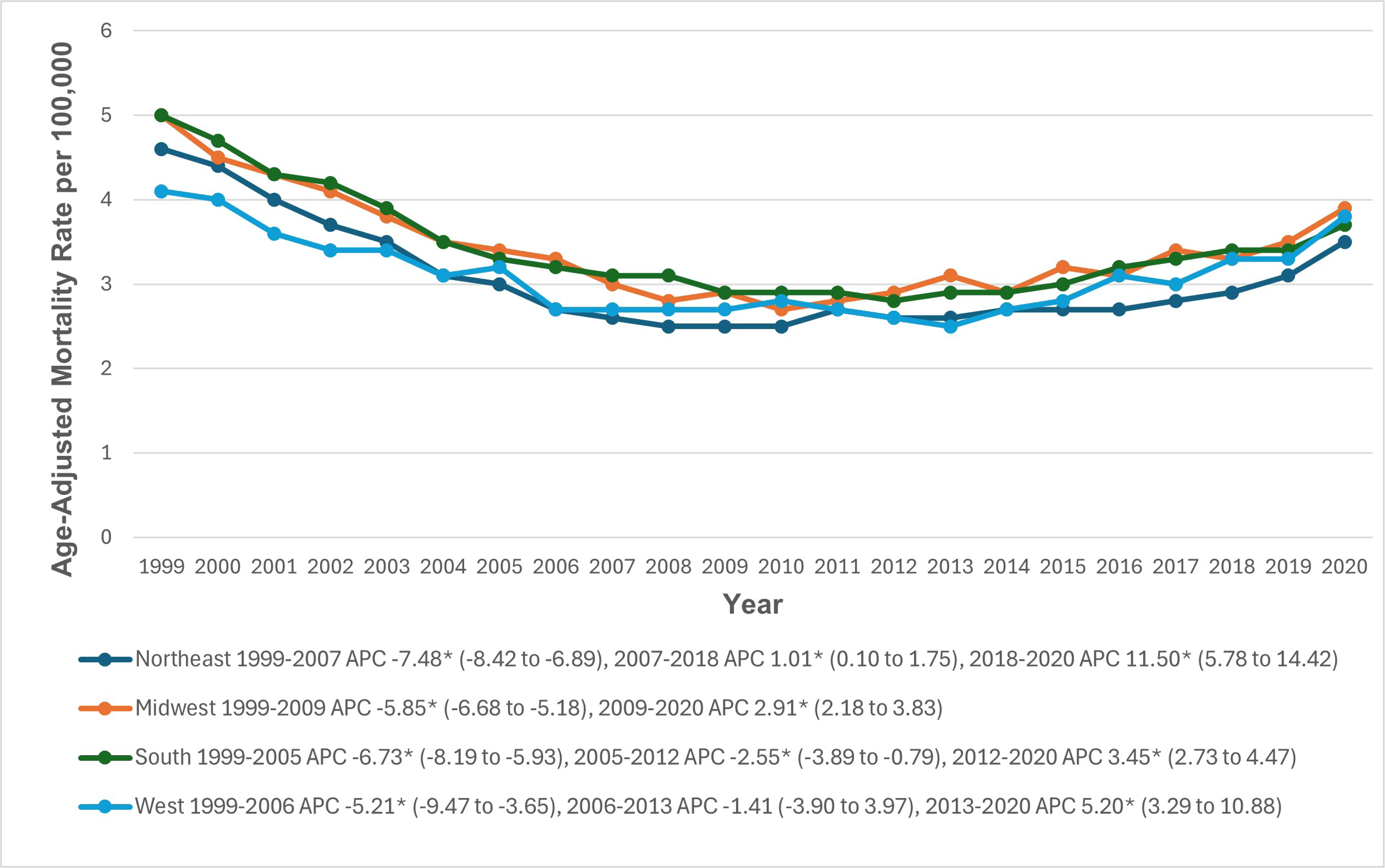
Paroxysmal Tachycardia-related Age-Adjusted Mortality Rates per 100,000, Stratified by Census Regions in Adults in the United States, 1999 to 2020 * Indicates that the annual percentage change (APC) is significantly different from zero at α = 0.05. AAMR = age-adjusted mortality rate.

### Urbanization

Throughout the study period, non-metropolitan areas had higher AAMRs for Paroxysmal tachycardia than metropolitan areas, with overall AAMRs of 3.6 (95% CI: 3.6 to 3.6) and 3.2 (95% CI: 3.2 to 3.2), respectively. Among the metropolitan group, AAMRs significantly decreased from 1999 to 2007 (APC: -6.31; 95% CI: -6.89 to -5.84), followed by a slight decrease from 2007 to 2014 (APC: -0.28; 95% CI: -1.23 to 0.69) and a significant increase till 2020 (APC: 4.37; 95% CI: 3.63 to 5.47). Among the non-metropolitan group, there was a significant decrease in AAMRs from 1999 to 2006 (APC: -5.50; 95% CI: -7.45 to -4.37), followed a slight decrease from 2006 to 2012 (APC: -1.56; 95% CI: -6.40 to 0.30). The AAMRs increased slightly from 2012 to 2018 (APC: 2.61; 95% CI: -1.35 to 4.14), followed by a significant increase till 2020 (APC: 8.96; 95% CI: 4.35 to 11.92). (Figure 6, Supplemental Table 3 and 9).

**Figure 6.**
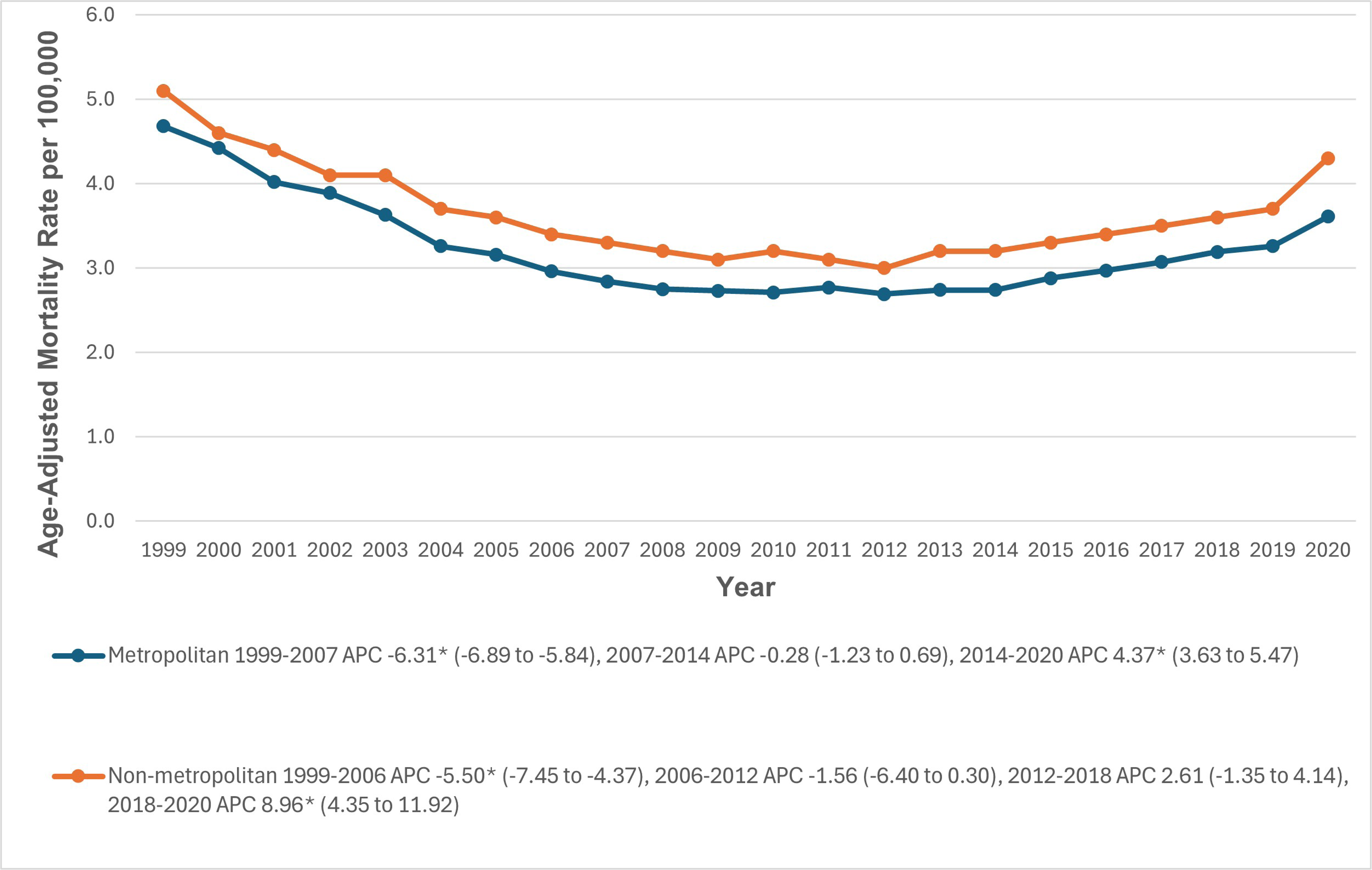
Paroxysmal Tachycardia-related Age-Adjusted Mortality Rates per 100,000 in Adults in the Metropolitan and Non-metropolitan areas in the United States, 1999 to 2020 * Indicates that the APC is significantly different from zero at α = 0.05. AAMR = age-adjusted mortality rate.

## Discussion

The current study reveals trends, gender, age, racial, and geographical differences in mortality due to paroxysmal tachycardia. Up to the authors’ best knowledge, this is the first study to investigate these trends from 1999 to 2020 in the US population. Data obtained from the Centers for Disease Control and Prevention’s WONDER database The data comprised 155,320 deaths caused by paroxysmal tachycardia from 1999 to 2020. During the period of 1999 to 2008, there was a steady decline in mortality due to paroxysmal tachycardia. However, it was followed by a steady incline in mortality rate, reaching 10008 deaths in 2020, which is higher than deaths recorded in 1999, which accounted for 8387 cases. This finding could be attributed to the fact that cardiac arrhythmias has been documented as a potential complication of COVID-19 (13).

The trends in gender and age differences noted in our study signal a critical area for further investigation and intervention. Men had higher AAMRs as compared to women. This variation may result from differences in the risk of ischemic heart disease and variations in the susceptibility of different genders to arrhythmias (14). Gender differences have been associated with certain claims relating to hormones, where it has been documented that testosterone lowers the calcium release facilitated by ryanodine receptors and the sodium/calcium exchange current (14). Due to the opposite action of estrogen, women may be more susceptible to provocative activity (15, 16). On the contrary, estrogen is also known to be cardioprotective, as indicated by the decrease of cardiac cell death in cardiomyopathies while testosterone stimulates the production of inflammatory substances (17).

Regarding age groups, older adults had the highest AAMRs due to paroxysmal tachycardia compared to the middle and young adult age groups, which could be attributed to older adults having a higher number of comorbidities placing them at increased risk. Notably, there has been an increase in AAMR among all age groups in the last decade, which could be explained by the COVID-19 pandemic, as arrhythmias are potential complications, and also by increased accessibility to healthcare and improved technology, which eases diagnosis.

In the context of racial differences, the most significant mortality due to paroxysmal tachycardia was noted in the NH Black or African American group, followed by the NH Indian American racial population. Racial differences are most likely due to socioeconomic and environmental factors (18). This gap may be linked to recognized discrepancies in healthcare access and resources encountered by these minority communities (19). Such could include income and insurance differences, decreased access to healthcare, and the prevalence of unmanaged comorbidities, which could all act as risk factors for paroxysmal tachycardia and arrhythmias. Future research must therefore focus on exploring socioeconomic factors which could contribute to potential disparities. Moreover, this is also reinforced by previous evidence, which highlights greater COVID-19 infection rates and deaths within these groups (20).

Furthermore, we noticed in our study that the AAMR of paroxysmal tachycardia was higher in the non-metropolitan region compared to the metropolitan region; however, overall increases in mortality rates were noticed in both regions in the last decade. Due to their limited access to healthcare services, non-metropolitan areas may have seen a rise in the mortality gap between urban and rural areas (21). While telehealth is not appropriate for every patient, a study by Soliman et al. showed that it can be used to manage primary and secondary cardiovascular disease in a large patient population (22). Telehealth platforms can be utilized to follow up patients with comorbidities and ensure they are managed to mitigate their risk of complications.

### Limitations

Our study has multiple areas of limitations. As the nature of the study is observational and retrospective, this study did not explore the hidden confounders such as income, education, exercise level, past smoking and alcohol use, and ease of access to medical care. Therefore, it is crucial to proceed cautiously with our findings and conduct future studies by including multiple data sources or methodologies to reproduce or verify the results. Secondly, the data included in our research was obtained from a single source (WONDER), which might not have included all aspects of arrhythmia in the US. Importantly, coding errors are conceivable because this mortality research relied on death certificates and the ICD-10 classification for the cause of death. Furthermore, we are unaware of the baseline comorbidities, CVD risk factors, mental health status, and socioeconomic situation, in addition to other medical issues. Finally, our study is based in the United States which limits its generalizability to other populations, and this highlights that further research is essential to understand the demographic differences.

### Future Prospects

Overall, this study reveals a significant mortality rate associated with paroxysmal tachycardia in the United States. By addressing possible contributory variables such as pandemic effects and racial and ethnic inequalities, healthcare providers and policymakers may devise comprehensive solutions to combat this concerning trend. Education and information should not only focus on identifying risk factors but also on modifying the socio-cultural contexts that lead to the same risk factors. In this setting, tight partnerships between policymakers, health care providers, and doctors are also crucial for ensuring a favorable cost-effectiveness ratio of health spending (23).

## Conclusions

Overall, our findings revealed that, despite a decrease in the rate of paroxysmal tachycardia-related mortality in the United States between 1999 and 2020, there is a rising mortality rate among males and NH Black or African Americans, particularly in non-metropolitan areas. This highlights that arrhythmia is becoming more prevalent in these populations.

Furthermore, the increased rate in paroxysmal tachycardia-related mortality last year shows the need for additional research into etiology, which would aid in the implementation of tailored therapies to improve care in a specific group. Despite significant advances in treatment and secondary prevention approaches, this study emphasizes demographic differences. These findings necessitate immediate public health actions to reduce these potential disparities.

## Data Availability

The data supporting the findings of this study are openly available in CDC Wonder database.

[https://wonder.cdc.gov/]

## Acknowledgements

The authors have no acknowledgements to declare.

## Abbreviations

(SVT): supraventricular tachycardia
(VT): ventricular tachycardia
(AAMRs): Age-adjusted mortality rates
(APC): Annual percentage change
(NH): Non-Hispanic

---

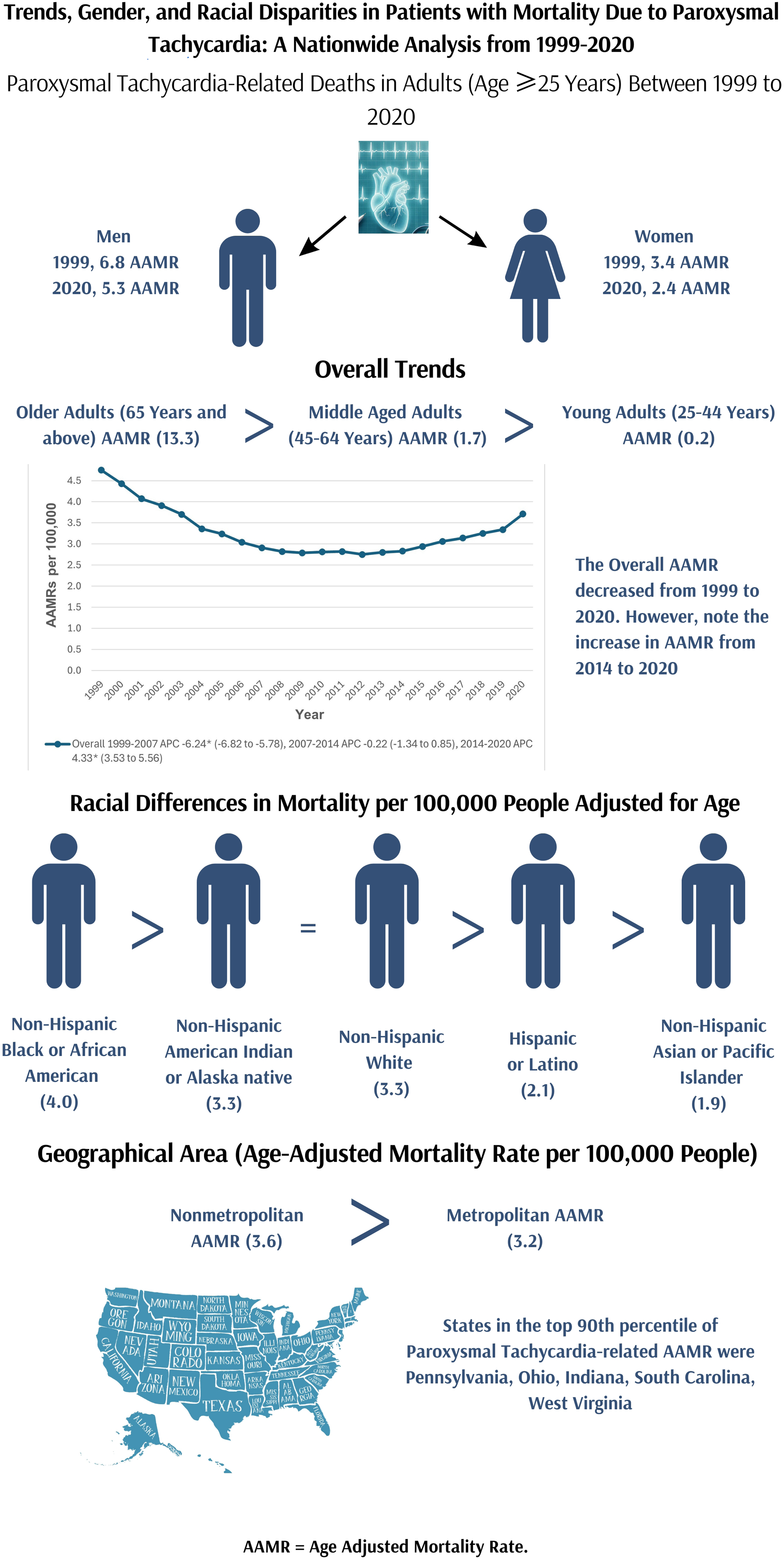

